# Presence of SARS-Coronavirus-2 in sewage

**DOI:** 10.1101/2020.03.29.20045880

**Authors:** Gertjan Medema, Leo Heijnen, Goffe Elsinga, Ronald Italiaander, Anke Brouwer

**Affiliations:** KWR Water Research Institute, Nieuwegein, The Netherlands

**Author notes:** Corresponding author contact Gertjan Medema, KWR Water Research Institute, Groningenhaven 7, 3433PE Nieuwegein, The Netherlands.

## Abstract

In the current COVID-19 pandemic, a significant proportion of cases shed SARS-Coronavirus-2 (SARS-CoV-2) with their faeces. To determine if SARS-CoV-2 is present in sewage during the emergence of COVID-19 in the Netherlands, sewage samples of 7 cities and the airport were tested using RT-PCR against three fragments of the nucleocapsid protein gene (N1-3) and one fragment of the envelope protein gene (E). No SARS-CoV-2 was detected in samples of February 6, three weeks before the first case was reported in the Netherlands on February 27. On March 5, the N1 fragment was detected in sewage of five sites. On March 15/16, the N1 fragment was detected in sewage of six sites, and the N3 and E fragment were detected at 5 and 4 sites respectively. This is the first report of detection of SARS-CoV-2 in sewage. The detection of the virus in sewage, even when the COVID-19 prevalence is low, indicates that sewage surveillance could be a sensitive tool to monitor the circulation of the virus in the population.

## Introduction

In December 2019, an outbreak of coronavirus respiratory disease (called COVID-19) initiated in Wuhan, China. The outbreak was caused by a new severe acute respiratory syndrome coronavirus 2 (SARS-CoV-2). The outbreak spread from Wuhan to other cities in China and many other countries. WHO declared a pandemic on March 11, 2020, when the disease was reported in 114 countries.^1^ The primary mode of transmission is via respiratory droplets that people cough, sneeze or exhale, and may also be spread via fomites.^2^ SARS-CoV-2 is 82% similar to SARS coronavirus that caused an outbreak in 2003. 16–73% of patients with SARS were reported to have diarrhea in addition to respiratory symptoms^3^, and transmission of SARS via water droplets from faeces via air ventilation systems in Amoy Gardens in Hong Kong was reported.^4^ Diarrhea is also reported in a significant proportion of the COVID-19 cases and recent reports show that SARS-CoV-2 has been detected in stool samples of COVID-19 cases.^5-9^ The shedding of SARS-CoV-2 was studied in a cluster of 9 cases and was 10^7^ RNA copies/g faeces one week after symptom onset and decreased to 10^3^ RNA copies/g three weeks after symptom onset.^10^ In stool samples with high RNA copies, viable SARS-CoV-2 was detected.^11^ Although it is unlikely that wastewater will become an important transmission pathway for coronaviruses like SARS-CoV-2^12^, increasing circulation of the virus in the population will increase the virus load into the sewer systems of our cities. It is important to collect information about the occurrence and fate of this new virus in sewage to understand if there is no risk to sewage workers, but also to determine if sewage surveillance^13^ could be used to monitor the circulation of SARS-CoV-2 in our communities, that could complement current clinical surveillance, which is limited to the COVID-19 patients with the most severe symptoms. Sewage surveillance could also serve as early warning of (re-)emergence of COVID-19 in cities, much like the sewage surveillance for poliovirus that has been used for this purpose.^14^ The objective of this investigation was to identify if SARS-CoV-2 is present in domestic wastewater of cities and a main airport during the early stages of the COVID-19 epidemic in the Netherlands.

## Methods

### Sewage samples

Before the onset of the epidemic in the Netherlands, wastewater treatment plants (WWTP) were selected that served 2 large and 3 medium sized cities and the main airport. The operators of the WWTP sampled a 24h flow-dependent composite sample of 250 mL that was stored at 4 °C during sampling. Samples were taken in 2020, on February 5, 6 and 7, 3 weeks before the first COVID-19 case was recognized by the health surveillance system in The Netherlands, on March 4 and 5 (38 resp. 82 reported COVID-19 cases in the Netherlands, total population 17.2 million) and March 15 and 16 (1135 resp. 1413 reported COVID-19 cases in the Netherlands). As the epidemic progressed, a WWTP (Tilburg) in one of the most affected areas was included in the sampling scheme.

### Sample processing

The samples were transported to the laboratory on melting ice and RNA was isolated on the day of sampling. Larger particles (debris, bacteria) were removed from the samples by pelleting using centrifugation at 4654xg for 30 mins without brake. A volume of 100-200 ml supernatant was filtered through Centricon^®^ Plus-70 centrifugal ultrafilter with a cut-off of 100 kDa (Millipore, Amsterdam, the Netherlands) by centrifugation at 1500xg for 15 minutes. The resulting concentrate was split in two parts for: 1) quantitative culturing for F-specific RNA phages and 2) for RNA extraction and RT-PCR.

Two procedures were used to extract RNA from the concentrated sewage samples. The samples of February 5, 6 and 7 and March 4 and 5 were processed using the RNeasy PowerMicrobiome Kit (Qiagen, Hilden, Germany) according to the manufacturers protocol. For practical reasons it was decided to use the magnetic extraction reagents of the Biomerieux Nuclisens kit (Biomerieux, Amersfoort, the Netherlands) in combination with the semi-automated KingFisher mL (Thermo Scientific, Bleiswijk, The Netherlands) purification system to extract RNA from the Centricon concentrates of the samples of March 15 and 16 as previously described.^15^ Elution of RNA was done in 100 µl elution buffer for both RNA extraction methods. The volume of sewage used for RNA extraction is shown in Table 1 for each of the samples.

**Table 1.**
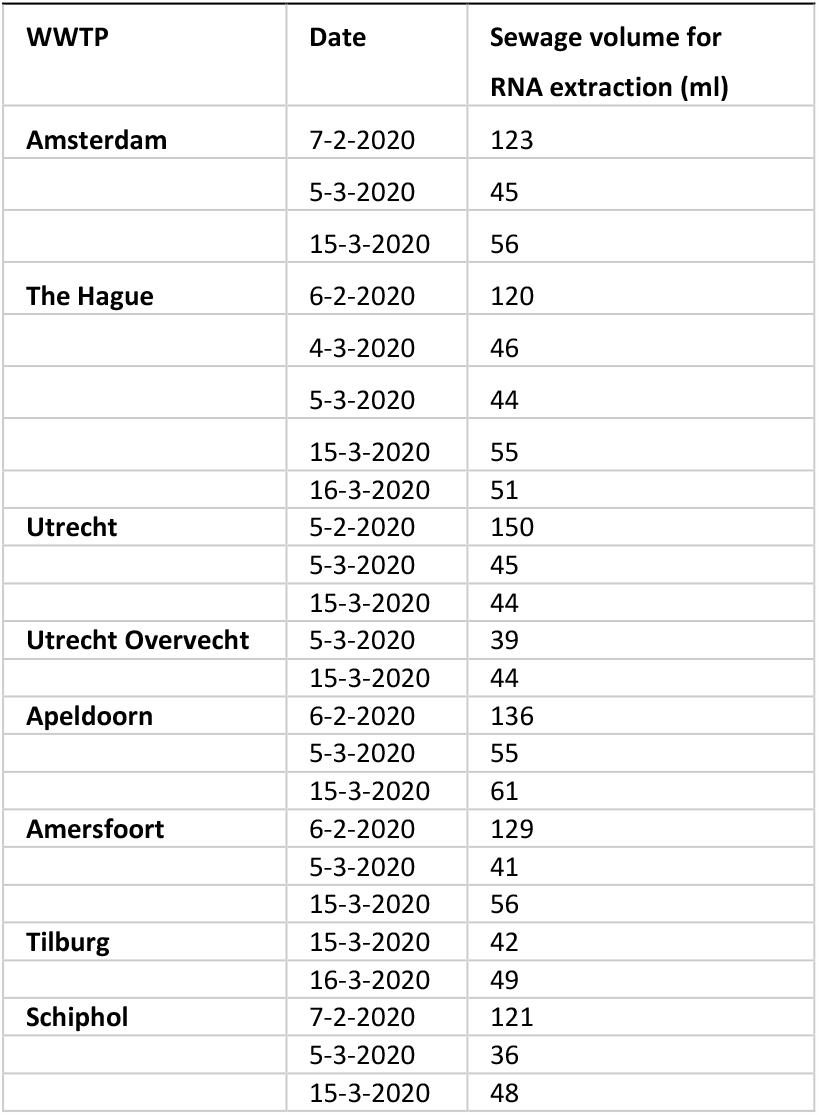
Sewage volumes used for RNA extraction

### Real-time RT-PCR

Primers/probe sets that were published by US CDC^16^ and a European study^17^ were used in this study (table 2). Four primer sets were selected (Table 2): the N1-N3 sets from CDC that each target a different region of the nucleocapsid (N) gene and the set against the envelope protein (E) gene from Corman et al.^17^, to include targets against two separate SARS-CoV-2 genes. The specificity of these primer/probe sets against other (respiratory) viruses, including human coronaviruses, was reported.^16,17^ Each reaction contained 5 µl RNA template, 4 µl of 5x EvoScript RNA Probes one-step RT-PCR reaction master mix (Roche Diagnostics, Almere, The Netherlands), different concentrations of primers and probes (table 2), 2 µl of 4 mg/ml BSA (Bovine Serum Albumin, Roche Diagnostics, Almere, The Netherlands) and the reaction volume was adjusted to a final volume of 20 µl with RT-PCR grade water (delivered with the Evoscript master mix). Thermal cycling reactions were carried out at 60 °C for 15 minutes, followed by 95 °C for 10 minutes and 45 cycles of 95 °C for 10 and 55 °C for 30 seconds on a CFX96 Touch Real-Time PCR Detection System (Bio-Rad Laboratories, Veenendaal, The Netherlands). Reactions were considered positive if the cycle threshold was below 40 cycles.^16^

**Table 1.**
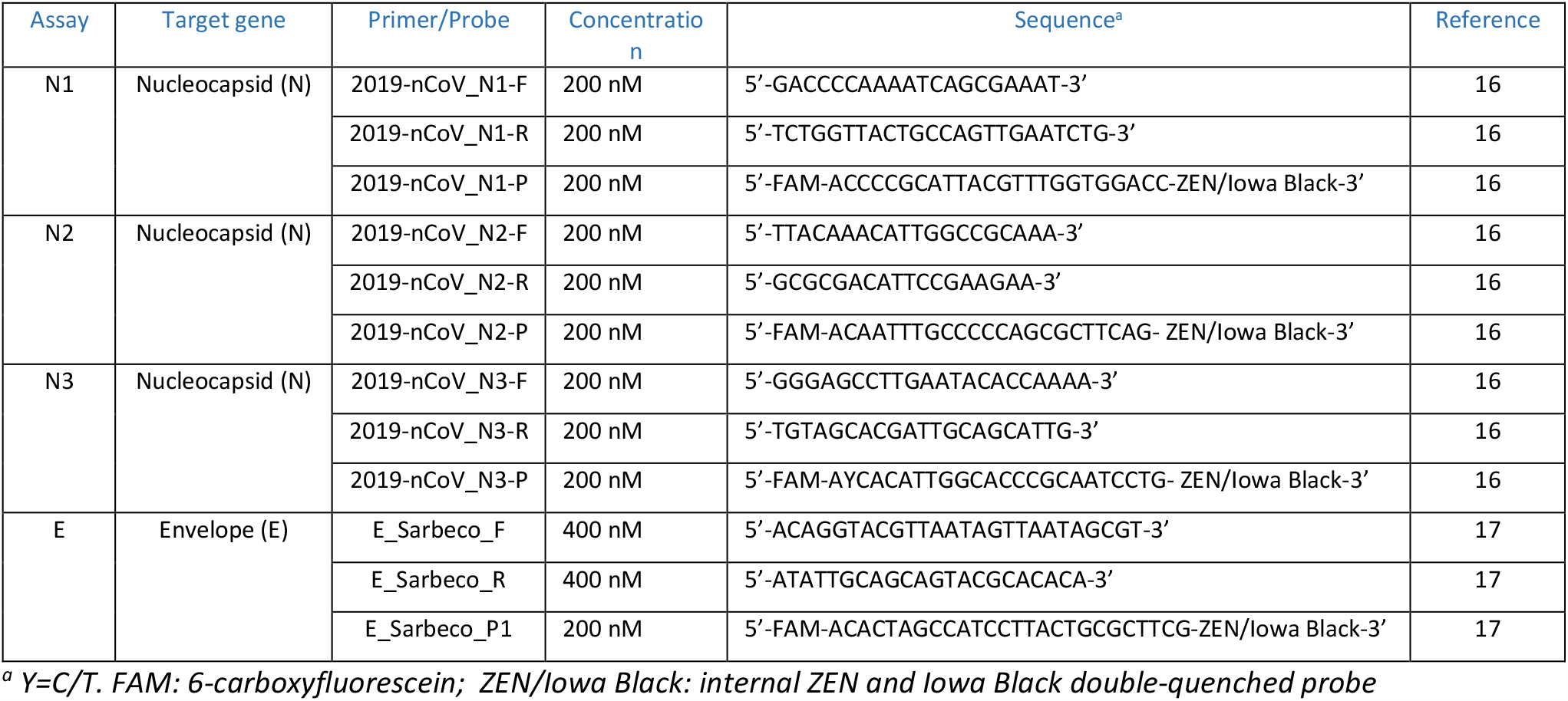
Primer-probe sets

### Controls

The concentration of F-specific RNA phages was measured according to ISO 10705 in wastewater before and after the purification and concentration steps, to determine the virus recovery. A previously described non-target RNA fragment^18^ was added to the lysed sewage concentrates as an internal control (IC). This IC-RNA consists of an in vitro transcribed RNA fragment with a length of 412 bases derived from dengue virus type 2 and was synthesized as previously described.^18^ RT-PCR analyses using primers IC-F (5′-ATGACAGCCACTCCTCCG-3′), IC-R (5′-GGAACGAACCAAACAGTCTTC-3′) and probe IC-P (5′-TexasRed-AGCAGAGACCCATTCCCTCAGAGC-BHQ-3′) were used to detect a dengue virus fragment (length: 149 bp) in order to control the performance of RNA-extraction, RT-PCR and to detect the presence of inhibitors. High-resolution automatic electrophoresis was performed on an Agilent 2100 Bioanalyzer using Agilent 1000 DNA Kit (Agilent, Santa Clara, USA) to analyse the length of the PCR-products.

## Results

The recovery of F-specific RNA phages by the purification and concentration steps was 73 ± 50% (n=16). Initial trials showed inhibition of the RT-PCR reactions, that could be reduced to a large extend by the addition of BSA to the reaction mixture. The results of the samples of February 6, 2020, three weeks before the first case was reported in The Netherlands on Feb 27, showed no positive signals for primer sets N1-3 and E (Table 3). The samples of March 4 and 5, one week into the epidemic with 38 and 82 COVID-19 cases reported through the health surveillance system, respectively^19^, showed a positive signal for the N1 primer/probe set in sewage samples of four of the 6 WWTP sampled. In sewage samples of March 15 and 16, 6 of the 7 WWTP showed a positive signal with N1, and in addition 5 WWTP also with N3 and 4 WWTP with the E primer/probe set. High throughput electrophoresis confirmed that the length of the PCR products match the length of the PCR target gene fragments. N1 appears to be the most sensitive primer/probe set of the sets tested here, followed by N3 and E for detection of SARS-CoV-2 in sewage. For clinical samples, the US FDA reported the sensitivity of the primer/probe sets of N1=N3>N2 on SARS-CoV-2 RNA,^20^ which partially matches with our results in sewage samples. The detection of fragments of two genes of SARS-CoV-2 in sewage of multiple WWTP and the temporal pattern of detection that aligns with the emergence of the epidemic in the Netherlands provide compelling evidence that SARS-CoV-2 is detected in sewage.

**Table 3.**
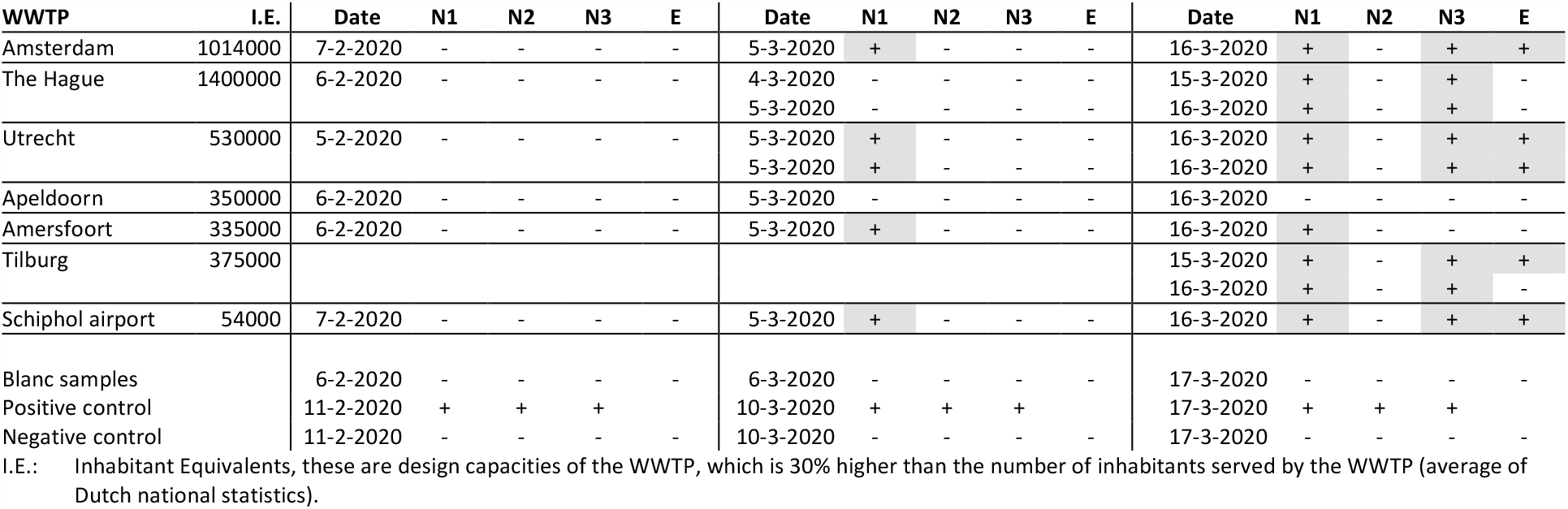
Results of screening of SARS-CoV-2 targets in 24h composite samples of incoming wastewater at different WWTP in The Netherlands 3 weeks before and approx. 1 and 2.5 weeks after the first COVID-19 case was reported in The Netherlands (February 27, 2020).

Looking for a RT-qPCR signal between 40 and 45 cycles showed signal in the March 16 samples also for the N2 primer/probe set, in the same six WWTP as set N1 was positive. In addition WWTP Apeldoorn yielded an increased signal of set N1 and E on March 16. Resampling these sites if the prevalence of COVID-19 increases in the catchment area of these WWTP will show if the signal with N2 and the other sets increase further.

There is no epidemiological signal or case reports in the Netherlands or other countries that are hit by the COVID-19 pandemic that sewage is a transmission route for SARS-CoV-2. Although SARS-CoV-2 is found with RT-PCR in a significant proportion of stool samples^5-10^, only two studies reported culture of infectious virus from stool.^11,21^ Therefore, sewage does not seem to be a transmission pathway of significance for SARS-CoV-2.^12^

### Sewage surveillance

To get an indication of the sensitivity of the monitoring of sewage, a proxy for the prevalence of COVID-19 in the cities served by the WWTP sampled was created using: 1. the number of COVID-19 cases on March 5 or 16, as reported for the cities that are served by each of the WWTP as numerator, and 2. the number of people served by each of the WWTP as denominator. The latter was estimated from the capacity (in inhabitant equivalents) of each of the WWTP. The prevalence at the airport could not be estimated, as the number of COVID-19 cases were not reported for this denominator. This yielded a rough estimate of the prevalence, since the service areas of WWTP do not precisely overlap with the city boundaries. Also, there was some delay in notification of cases by the local health authorities to the national surveillance system which could mean that the actual prevalence was slightly higher than reported. Moreover, the reported prevalence are the data from cases with COVID-19 as confirmed by laboratory diagnosis; a significant proportion of COVID-19 goes undetected, since people with mild symptoms are not tested. A study among healthcare workers in two hospitals in the Netherlands indicated that SARS-CoV-2 was already circulating undetected in the community prior to February 27, when the first COVID-19 case was reported, suggesting that there is a high prevalence of mild COVID-19 in the community.^22^ The detection of N1 in WWTP Amersfoort on March 5, when no cases were reported in Amersfoort, also suggests virus circulation in the population before COVID-19 cases are reported through the health surveillance system. Comparing Tables 3 and 4 shows that the N1 primer/probe set started to produce a signal in sewage samples when the observed COVID-19 prevalence was around or even below 1.0 case in 100,000 people and the N3 and E set started to yield positive signals when the observed prevalence was 3.5 case per 100,000 people or more, although not consistently, since sewage from WWTP Amersfoort did not yield positive results with set N3 and E. Given the roughness of the prevalence estimates, these numbers are indicative, but do indicate that sewage surveillance with the method used in this study is sensitive. However, reliable quantification of SARS-CoV-2 with RT-qPCR in sewage will be required to make reliable surveillance feasible. Therefore, the development of controls to consistently monitor coronavirus recovery and to measure viral RNA yield and check for RT-PCR inhibition is of great importance. The analyses of an added quantified suspension of another human coronavirus (such as 229E)^23^ to the sewage samples can potentially be used as an easy control to make reliable quantification possible. Also, digital droplet PCR could aid in the quantification of SARS-CoV-2 in water, as shown for other RNA viruses.^24^ The detection of the virus in sewage, even when the COVID-19 prevalence is low, indicates that sewage surveillance could be used to monitor the circulation of the virus in the population and as early warning tool for increased circulation in the coming winter or unaffected populations.

**Table 4.** Rough estimate of the observed prevalence of COVID-19 (number of infected persons per 100.000 people) in the community served by the WWTP on February 6, March 5 and 16, 2020.

| WWTP | Feb 27 | Mar 5 | Mar 16 |
| --- | --- | --- | --- |
| Amsterdam | 0,0 | 0,3 | 5.5 |
| The Hague area | 0,0 | 0,3 | 3.5 |
| Utrecht | 0,0 | 1,6 | 12.9 |
| Apeldoorn | 0,0 | 0,0 | 2.9 |
| Amersfoort | 0,0 | 0,0 | 7.7 |
| Schiphol | 0,0 | nd | nd |
| Tilburg | 0,0 | 0.9 | 32.0 |

## Data Availability

All data and methods are reported in the manuscript

## Acknowledgements

The authors are very grateful for the rapid assistance of the Water Authorities and WWTP operators in the Netherlands who organised the sampling and provided the sewage samples. We are also grateful to Meindert de Graaf for sample transport. This study was financed by KWR Water Research Institute.

## References

1 https://www.who.int/dg/speeches/detail/who-director-general-s-opening-remarks-at-the-media-briefing-on-covid-1911-march-2020 Last accessed Mar 23, 2020

2 https://www.ecdc.europa.eu/en/novel-coronavirus-china/questions-answers Last accessed Mar 23, 2020

3. WHO issues consensus document on the epidemiology of SARS. Wkly Epidemiol Rec 2003 78: 373–75.

4. McKinney KR, Gong YY, Lewis TG. Environmental transmission of SARS at Amoy Gardens. J Environ Health 2006 68(9):26–30.

5. Wang D, Hu B, Hu C, Zhu F, Liu X, Zhang J, Wang B, Xiang H, Cheng Z, Xiong Y, Zhao Y, Li Y, Wang X, Peng Z. Clinical Characteristics of 138 Hospitalized Patients With 2019 Novel Coronavirus-Infected Pneumonia in Wuhan, China. JAMA 2020 Feb 7. doi: 10.1001/jama.2020.1585.

6. Chen N, Zhou M, Dong X, Qu J, Gong F, Han Y, Qiu Y, Wang J, Liu Y, Wei Y, Xia J, Yu T, Zhang X, Zhang L. Epidemiological and clinical characteristics of 99 cases of 2019 novel coronavirus pneumonia in Wuhan, China: a descriptive study. Lancet 2020 395(10223):507–513.

7. Holshue ML, DeBolt C, Lindquist S, Lofy KH, Wiesman J, Bruce H, Spitters C, Ericson K, Wilkerson S, Tural A, Diaz G, Cohn A, Fox L, Patel A, Gerber SI, Kim L, Tong S, Lu X, Lindstrom S, Pallansch MA, Weldon WC, Biggs HM, Uyeki TM, Pillai SK; Washington State 2019-nCoV Case Investigation Team. First Case of 2019 Novel Coronavirus in the United States. N Engl J Med 2020 382(10):929–936.

8. Xiao F, Tang M, Zheng X, Liu Y, Li X, Shan H. Evidence for gastrointestinal infection of SARS-CoV-2. Gastroenterology 2020 Mar 3. pii: S0016-5085(20)30282-1. doi: 10.1053/j.gastro.2020.02.055.

9. Xu Y, Li X, Zhu B, Liang H, Fang C, Gong Y, Guo Q, Sun X, Zhao D, Shen J, Zhang H, Liu H, Xia H, Tang J, Zhang K, Gong S. Characteristics of pediatric SARS-CoV-2 infection and potential evidence for persistent fecal viral shedding. Nat Med 2020 https://doi.org/10.1038/s41591-020-0817-4

10. Woelfel R, Corman V, Guggemos W, Seilmaier M, Zange S, Mueller MA, Niemeyer D, Vollmar P, Rothe C, Hoelscher M, Bleicker T, Bruenink S, Schneider J, Ehmann R, Zwirglmaier K, Drosten C, Wendtner C. Clinical presentation and virological assessment of hospitalized cases of coronavirus disease 2019 in a travel-associated transmission cluster. medRxiv 2020.03.05.20030502; doi: https://doi.org/10.1101/2020.03.05.20030502

11. Wang W, Xu Y, Gao R, et al. Detection of SARS-CoV-2 in Different Types of Clinical Specimens. JAMA. Published online March 11, 2020. doi:10.1001/jama.2020.3786

12. WHO/UNICEF Water, sanitation, hygiene and waste management for the COVID-19 virus. Technical brief, 3 March 2020.

13. Hellmér M, Paxéus N, Magnius L, Enache L, Arnholm B, Johansson A, Bergström T, Norder H. Detection of pathogenic viruses in sewage provided early warnings of hepatitis A virus and norovirus outbreaks. Appl Environ Microbiol 2014 80(21):6771–81.

14. Hovi T, Shulman LM, van der Avoort H, Deshpande J, Roivainen M, de Gourville EM. Role of environmental poliovirus surveillance in global polio eradication and beyond. Epidemiol Infect 2012 140(1):1–13.

15. Heijnen L, Medema G. Surveillance of influenza A and the pandemic influenza A (H1N1) 2009 in sewage and surface water in the Netherlands. J Water Health 2011 9(3):434–42.

16. 2019-Novel coronavirus (2019-nCoV) real-time rRT-PCR panel primers and probes. US Centers for Disease Control and Prevention. Last accessed Mar 24, 2020.

17. Corman VM, Landt O, Kaiser M, Molenkamp R, Meijer A, Chu DKW, Bleicker T, Brünink S, Schneider J, Schmidt ML, Mulders Dgjc, Haagmans BL, van der Veer B, van den Brink S, Wijsman L, Goderski G, Romette JL, Ellis J, Zambon M, Peiris M, Goossens H, Reusken C, Koopmans MPG, Drosten C. Detection of 2019 novel coronavirus (2019-nCoV) by real-time RT-PCR. Euro Surveill 2020 Jan;25(3):2000045. doi: 10.2807/1560-7917.

18. Stinear T, Matusan A, Hines K, Sandery M. Detection of a single viable Cryptosporidium parvum oocyst in environmental water concentrates by reverse transcription-PCR. Appl Environ Microbiol. 1996 62(9):3385-90. Erratum in: Appl Environ Microbiol 1997 63(2):815.

19. https://www.fda.gov/media/136231/download Last accessed Mar 26, 2020.

20. https://www.rivm.nl/nieuws/actuele-informatie-over-coronavirus Last accessed Mar 16, 2020.

21. Yong Zhang Y, Chen C, Zhu S, Shu C, Wang D, Song J, Song Y, Zhen W, Feng Z, Wu G, Xu J, Xu W. Isolation of 2019-nCoV from a Stool Specimen of a Laboratory-Confirmed Case of the Coronavirus Disease 2019 (COVID-19)[J]. China CDC Weekly 2020, 2(8): 123–124

22. Kluytmans M, Buiting A, Pas S, Bentvelsen R, van den Bijllaardt W, van Oudheusden A, van Rijen M, Verweij L, Koopmans M, Kluytmans J. SARS-CoV-2 infection in 86 healthcare workers in two Dutch hospitals in March 2020. medRxiv 2020.03.23.20041913; doi: https://doi.org/10.1101/2020.03.23.20041913

23. Gundy PM, Gerba CP, Pepper IL. Survival of Coronaviruses in Water and Wastewater. Food Environ Virol 2009 1(1):10.

24. Rački N, Morisset D, Gutierrez-Aguirre I, Ravnikar M. One-step RT-droplet digital PCR: a breakthrough in the quantification of waterborne RNA viruses. Anal Bioanal Chem 2014 406(3):661–7.

